# Public’s Knowledge and Practices regarding COVID-19: A cross-sectional survey from Pakistan

**DOI:** 10.1101/2020.06.01.20119404

**Authors:** Muhmmad Saqlain, Ali Ahmed, Aqsa Gulzar, Sahar Naz, Muhammad Muddasir Munir, Zaheer Ahmed, Sohail Kamran

## Abstract

Effective mitigation of coronavirus diseases (COVID-19) pandemic required true adoption of precautionary measures by the masses, that primarily depends upon their knowledge and practices behaviors. The current study aimed to evaluate the knowledge; practices of Pakistani residents regarding COVID-19 and factors associated with good knowledge and positive practices. A cross-sectional online survey was conducted from 15-April 2020 to 20 May 2020 among 689 Pakistanis by using a validated self-administered questionnaire. Regression analysis was applied to find potential predictor of knowledge and practices. Of 689 participants, more than half had good knowledge; 80% had good practices regarding COVID-19 and majority seek knowledge from social media. Knowledge was significantly higher (OR>1.00, *p*< 0.05) among educated and higher income participants. Positive practices were significantly (OR>1.00, *p*< 0.05) related to the older age (≥50 years), higher education, higher income and good knowledge regarding COVID-19. The study concluded that Pakistani residents had good knowledge and practices towards COVID-19 yet there are gaps in specific aspects of knowledge, and practice that should be focused in future awareness and educational campaigns. The study recommends the ministry of health authorities to promote all precautionary and preventive measures of COVID-19-consisting of a better-organized approach-to all strata of society: less privileged people, older ones and less educated people, in order to have equilibrium of knowledge about COVID-19; hence effective implementation of precautionary measures.

## Introduction

On 11 March 2020, the World Health Organization (WHO) declared the coronavirus disease (COVID-19) outbreak as a pandemic, with the spread of disease in 114 countries and more than 4000 deaths [1]. On 31 May 2020, there were 6,175,290 cases of COVID-19, with 371,228 confirmed deaths in 213 countries and territories of the world [2].

Pakistan, a developing nation, ranked 5th among the world’s populous countries to be considered the new COVID-19 hotspot with a fragmented health care system [3]. As far as COVID-19 is concerned, Pakistan has a uniquely challenging situation due to its vulnerable geographical location, as it shares borders with China and Iran due to poor screening capacity, leading to delayed implementation of preventive measures [4]. Pakistan’s first case of COVID-19 was reported on 26 February in Karachi, having a travel history of Iran [4]. By 31 May 2020, Pakistan has 68,270 laboratory-confirmed cases and 1483 COVID-19 associated deaths. Sindh (n = 27360) has the highest number of cases followed by Punjab (n = 25056), Khyber Pakhtunkhwa (KP) (n = 9540), Baluchistan (n = 4193), Islamabad (n = 2418), Gilgit Baltistan (678), and Azad Jammu Kashmir (n = 251) [5].

COVID-19 transmission has been confirmed to occur from human to human, and the virus is thought to spread through respiratory droplets when a person coughs or sneezes [6]. The most common symptoms are fever, shortness of breath, tiredness, cough and sore throat. Other rare symptoms include nasal congestion, runny nose, aches and diarrhea [6]. Some people have experienced a loss of taste or smell [7]. Older adults and people with underlying diseases like hypertension, diabetes, asthma, or cardiovascular disease are at risk of developing infection [7].

There are no recommendable effective drugs and vaccines to control the disease [8]. The best way to prevent infection is to avoid exposure to the virus by taking precautions to wash hands regularly with soap and water for at least 20 seconds, cover the nose and mouth with tissue or flexed elbow while coughing or sneezing, avoid close contact (1 meter or 3 feet) with people who are ill, avoid touching eyes, nose or mouth with dirty hands, stay home and self-isolate from others in case of feeling unwell [8, 9].

The ultimate impact of the COVID-19 pandemic outbreak is unclear at the moment, but more adherence to public health organizations’ suggestions will keep healthcare systems from being overwhelmed [10]. It is, therefore, imperative that the community be equipped with empirically precise knowledge and tools to address and cope with the impact of COVID-19 effectively. Currently, the COVID-19 is a health emergency and needs individual cooperation to let halt transmission by following given guidelines, government orders, and a range of preventive measures. Data suggest that knowledge, awareness, perception, and attitude of the general public regarding disease play an essential role in the control and management of disease as observed in epidemics of SARS and MERS [10, 11]. Poor knowledge of the general public regarding preventive measures as avoiding crowded areas, wearing masks, properly washing hands, and maintaining social distance also pose a significant gap in control of the spread of COVID 19 [12]. Deeming public awareness to be crucial in preventing the spread of COVID–19, which otherwise lacks effective treatment and preventive measures, vast public awareness campaigns are critical in the fight against it.

The Pakistani National Institute of Health (NIH) has played a vital role in designing and circulating protocols regarding COVID-19 transmission and prevention, as well as launching public awareness campaigns [13]. However, final success depends upon the adherence of people to guidelines and preventive measures that are strongly linked to their understanding and awareness towards disease. Therefore, there is an utmost need for the awareness level of the population in response to the outbreak of COVID-19. Survey highlighting awareness level is useful to get information regarding public health education, response and recovery efforts, and social mobilization [11, 14]. Data from the study is pivotal for policy development and public health implementation to respond to the outbreak shortly and consistently quickly.

In context of the explanation above, the current study aimed to evaluate the current level of awareness regarding transmission, symptoms, and preventive measures of COVID-19 among the general population in Pakistan. Additionally, this study will provide a snapshot of the extent of precautionary measures practiced by the Pakistani population.

## Methods

### Study design

A cross-sectional survey-based study was conducted during the month of April 2020, days of strict lockdown to implement social distancing to avoid the spread of the pandemic. The investigators opted for an online data collection method because it was not possible to carry out a population-based survey in this critical/censorious situation.

### Sampling, study population and data collection method

The sample size calculated by Raosoft was 583, assuming a response rate of 50%, confidence interval (CI) 95%, Z as 1.96, and margin of error d as 4%. Considering, an additional 20% (n = 116) for any error in questionnaire filling, a final sample size of 699 will be required. The survey was started on 15 April 2020, and response acceptance was closed on 20-May 2020, when required sample size was achieved. Pakistani citizens 16 years of age or older have been selected as a study population and agreed to fill out the data form/questionnaire.

The questionnaire was designed on google forms and the generated link was shared with the WhatsApp groups. Link was also shared personally with the contact list of investigators. Respondents from other provinces were also eligible to participate if they were willing to complete the questionnaire.

### Measure

A survey instrument was designed based on substantial literature analysis [14–16], material related to emerging respiratory diseases including COVID-19 by WHO [8] and guidelines issued by NIH, Islamabad Pakistan [13]. After the preliminary draft questionnaire was drawn up, it was validated in two stages. In the first place, the study tool was discussed with pharmacy and medical researchers and professionals to give their expert opinion on its simplicity, relativity and relevance. Secondly, a pilot study was conducted by selecting a small sample (n = 60) to make the questionnaire simpler and more comprehensive. The questionnaire was amended on the basis of the suggestions made by the participants and its consistency with the published literature. After a thorough discussion, the authors finalized the questionnaire and then distributed it to the participants for their response. The coefficient of reliability was calculated using SPSS v.20 and Cronbach’s alpha value. It was found to be 0.77. The data from the pilot study were not included in the final analysis.

The questionnaire included questions on the assessment of demographics, the source of information, knowledge, and practice of COVID-19. The demographic characteristics included gender, age, marital status, monthly income, residence, employment status and education. One item was regarding the source of information about COVID-19. Awareness section comprised of

20 items; regarding aetiology (2-items), symptoms (7-items), risk group (1-item), transmission (6-items), treatment (2-items) and precautions/preventions (2-items). Each question was responded as Yes, No, and I don’t know. The correct answer was marked as 1 while wrong answer was marked as 0. Total score ranges from 0–14, and a cut off level of ≤15 was set for poor knowledge and ≥16 (More than 75%) for good knowledge.

The practice section included 6 items related to the use of face mask, and implementation of other precautionary measures were included in the practice section. Each item was responded as yes (1-point), No (0-point), and sometimes (0-point). Practice items total score ranged as 0–6, where 5–6 score was considered as good practice, and a score of 1–4 indicated poor practice of preventive measures for COVID-19.

### Ethics

The study was performed following the declaration of Helsinki. Due to lockdown, universities were closed, hence study protocol was approved from the Hospital board (767/THQ/HR). The study questionnaire contained a consent portion that stated purpose, nature of the survey, study objectives, volunteer participation, declaration of confidentiality, and anonymity.

### Statistical analysis

Data were entered in Microsoft Excel and later imported into SPSS V.21 for statistical analysis. Numerical variables were measured as mean and standard deviations, while categorical variables were expressed as frequencies and percentages. Inferential statistics were applied depending upon the nature of data and variables. Chi-square tests were used to find differences in knowledge groups (good vs. poor) and practice (good vs. poor) by demographic characteristics. Binary logistic regression models have been used to identify possible determinants of good knowledge and practice, both unadjusted and adjusted (adjusted for age, gender and other demographic variables). Results were expressed as crude odds ratio (COR), and adjusted odds ratio (ARO) accompanied by 95% confidence interval (CI). A p-value of less than 0.05 will be considered significant in all tests.

## Results

Out of the total 699 responses collected, 10 questionnaires were excluded due to missing information, and 689 responses were analyzed in the final analysis. The majority of participants were male (62.6%, n = 431), 64.3% (n = 443) aged less than 30 years, and less than half (47.6%, n = 328) of participants were married. More than half (53.3%, n = 367) of respondent had monthly income of PKR 0 – 24,999, 76.8% (n = 529) were unemployed, and 65% (n = 448) participants had education of 13 years or more. (Table 1).

**Table 1.**
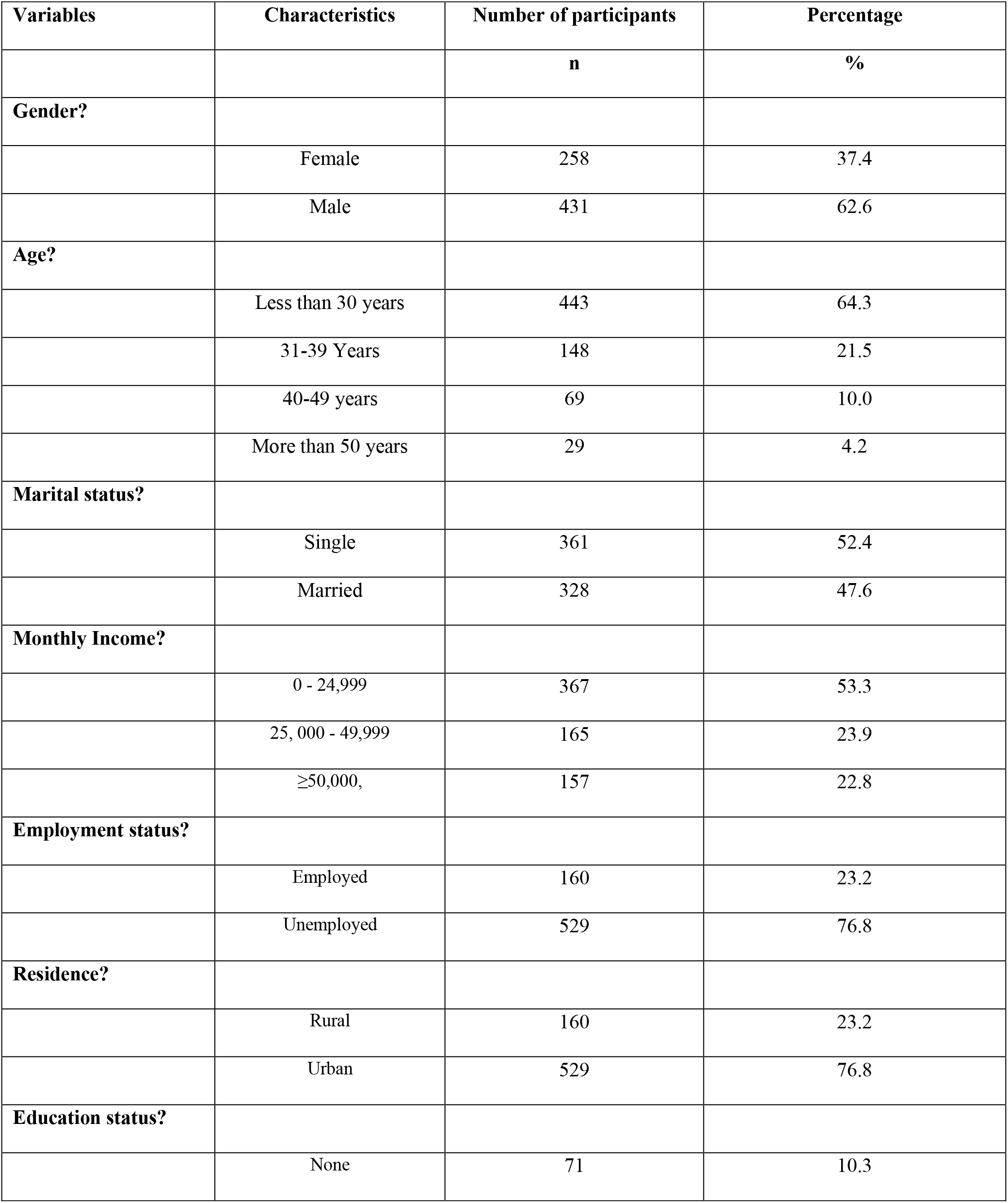
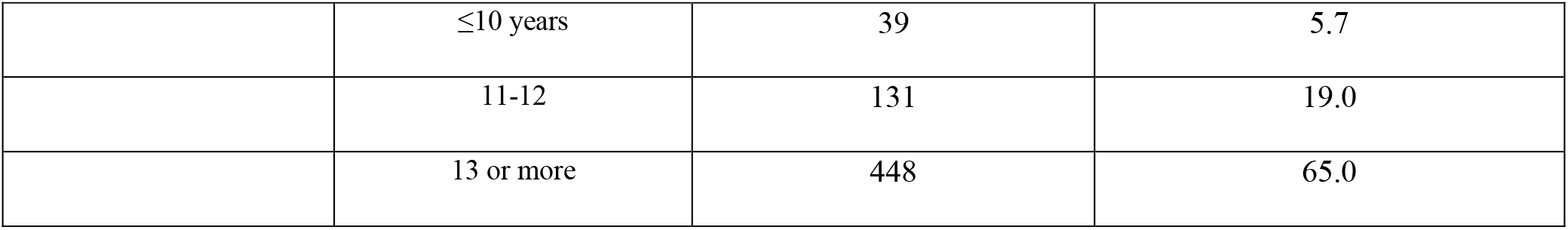
Demographic characteristics of study population (N = 689)

Fig. 1 provides a summary of the information sources utilized by respondents. Majority of participants used social media (66.62%, n = 459) as a source to seek information regarding COVID-19 followed by television/radio (62.99%, n = 434) and friends (25.54%, n = 176).

**Fig. 1.**
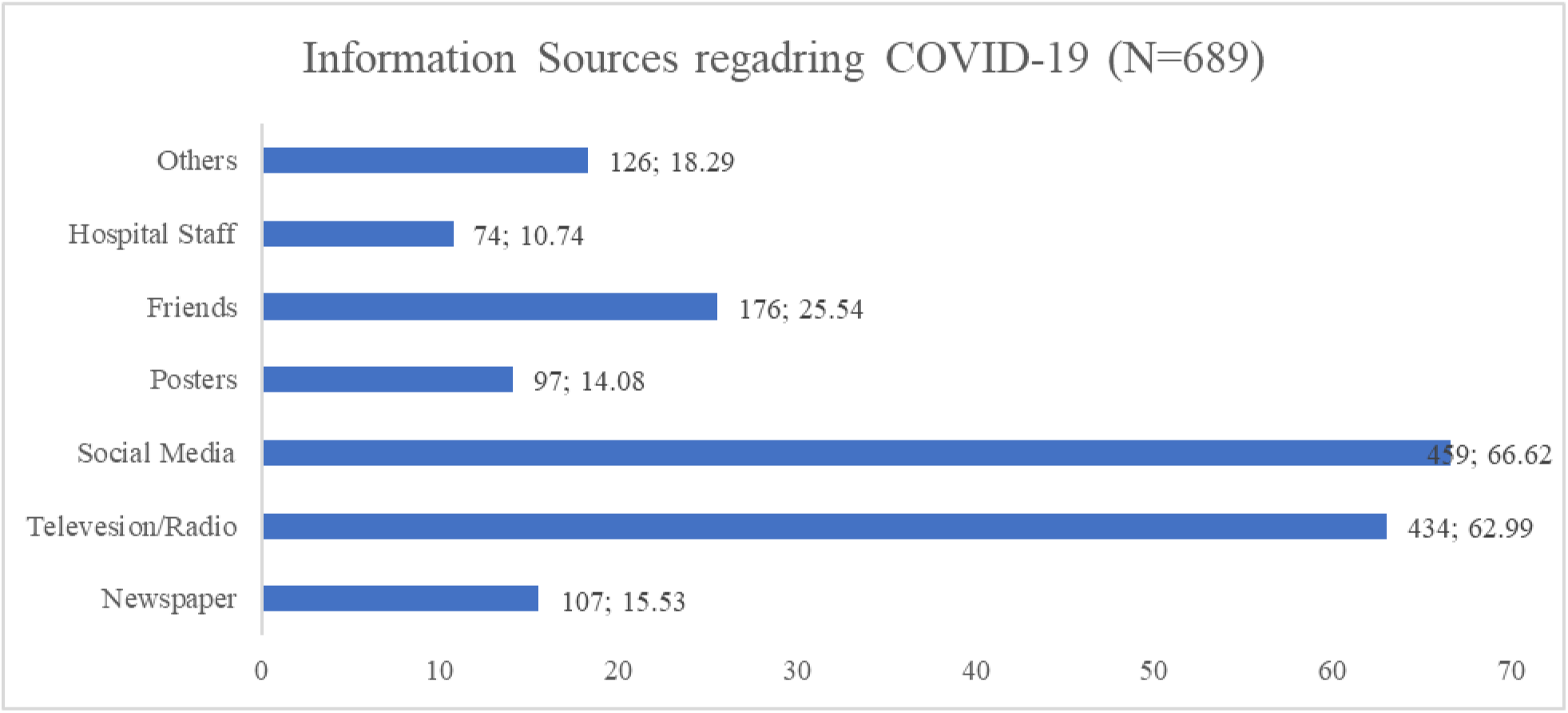
Information sources utilized by general public in Pakistan

### Knowledge regarding COVID-19

Fig. 2 summarizes the responses of participants for knowledge items of the questionnaire. Mixed responses were obtained regarding 20 items. Majority of participants had heard about disease, and 93.9 % (n = 647) know that virus is the causative agent of COVID-19. Response to questions regarding transmission of disease indicated 93.3% (n = 643) participants correctly identified that virus can transmit trough droplets, 97.2% (n = 670) subjects were well aware that infection can be transmitted by shaking hands, and 98.3% (n = 677) respondents had correct knowledge regarding the transmission of the virus from person to person. On the other hand, 53.3% and 47.8% of individuals respectively didn’t know that viruses can be transmitted from bats or other animals to humans.

**Figure 2.**
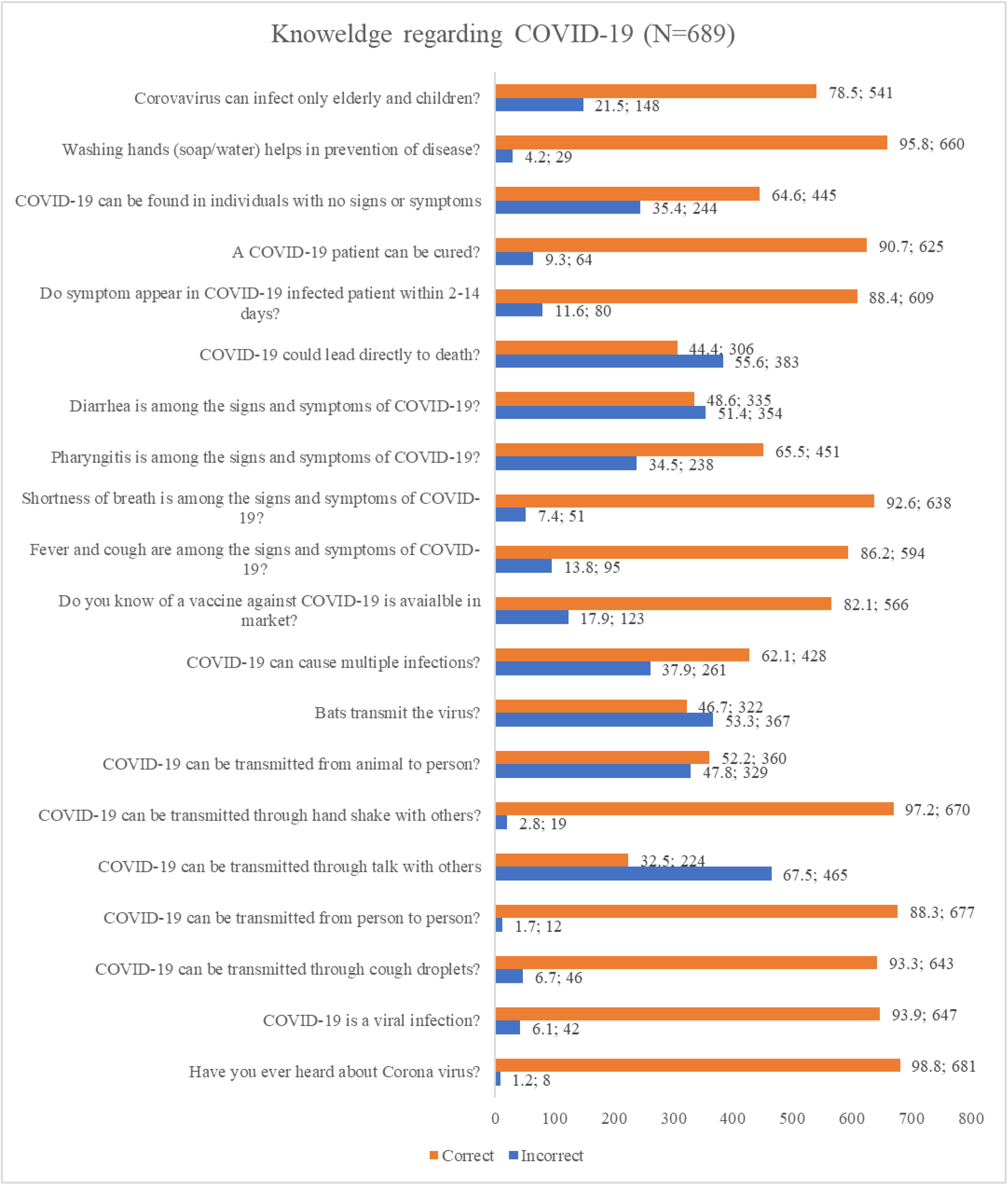
Public’s Knowledge regarding COID-19

More than 80% of respondents correctly know that fever, cough, and shortness of breath are the symptoms of COVID-19, while more than half (51.4%, n = 354) of participants had incorrect knowledge that diarrhea is a symptom of COVID-19. When the question asked regarding fatality of disease, 55.6% (n = 383) subjects respond that COVID-19 directly leads to death, 35.4% (n = 244) respondent didn’t know that COVID-19 can be found in a person with no signs and symptoms, and 21.5% (n = 148) reported that virus could affect only elderly and children.

### Practices regarding COVID-19

Fig. 3 summarizes the practices of the general population regarding COVID-19. All the participants respond to all 6 items regarding COVID0–19. The majority of respondents had good practice regarding each item with the highest practice showed towards washing of hands with soap or cleaning with sanitizers (94%, n = 648), and a similar proportion (93.8%, n = 646) of participants revealed that they avoid going in a crowded place. A lower percentage of good practice was observed among the general population to wear a face mask (79.8%, n = 550).

**Figure 3.**
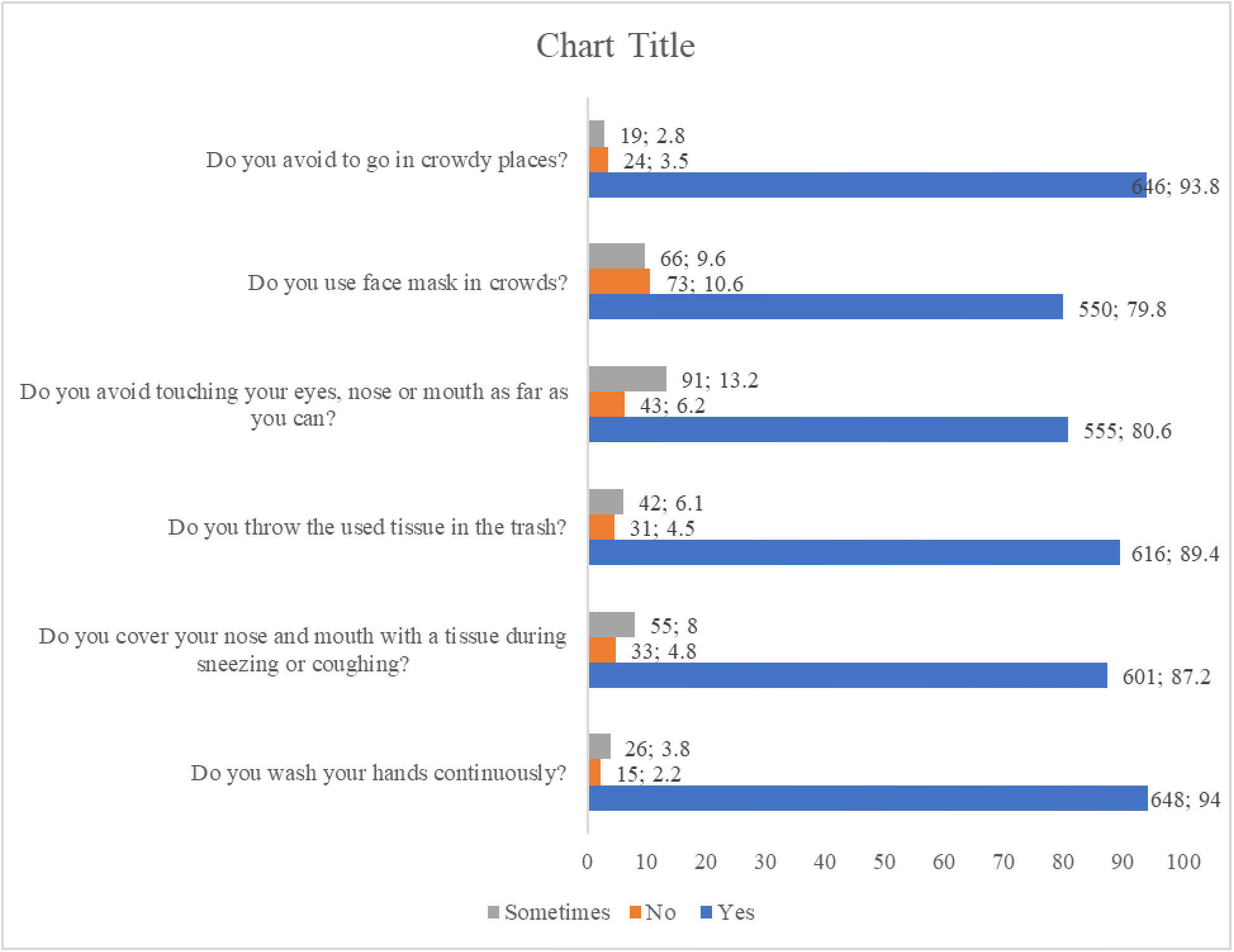
Public’s Knowledge regarding COID-19

### Difference in knowledge and practice status by demographics

More than half of participants had good knowledge (51.81%, n = 357), while 332 (48.19%) individuals had poor knowledge regarding COVID-19. Chi-square tests were applied to find differences in knowledge status by sample characteristics. Knowledge status was significantly differed by monthly income as participants with higher income have good knowledge compared to lower-income counterparts (**χ^2^** = 25.85, *p* < 0.001). The findings showed that knowledge status differed significantly from marital status (**χ^2^** = 4.606, *p* = 0.032), to employment status (**χ^2^** = 4.968, *p* = 0.026). Similarly, the majority of participants with higher education had good knowledge compared to less-educated counterparts (**χ^2^** = 24.07, *p*< 0.001) while the status of practice did not differ significantly in terms of gender, age, and residence (*p*>0.05).

Findings indicated that 80.55% (n = 555) participants had good practice in following precautionary measures regarding COVID-19. Chi-square analysis revealed that participant’s practices regarding COVID-19 were significantly differed by monthly-income (**χ^2^** = 8.979, *p* = 0.011), residence (**χ^2^** = 6.154, *p* = 0.013), and education status (**χ^2^** = 16.716, *p* = 0.001). While the status of the practice did not differ significantly in terms of gender, age, marital status, and employment status (*p*>0.05).

### Binary logistic regression analysis for factors associated with good knowledge and practice

Adjusted and un-adjusted regression analysis was applied to find possible predictors of good knowledge and practice among the general population in Pakistan. The regression model adjusted for all independent variables showed that participants with higher monthly income (≥50,000) had higher odds (AOR: 2.133, 95% CI: 1.394–3.867, *p*< 0.001) of good knowledge compared to the reference category. Similarly, participants who have an education of 13 years or more had higher odds (AOR: 1.501, 95% CI: 0.858–2.627, *p* = 0.048) of good knowledge compared to less-educated counterparts. (Table 3)

**Table 2.**
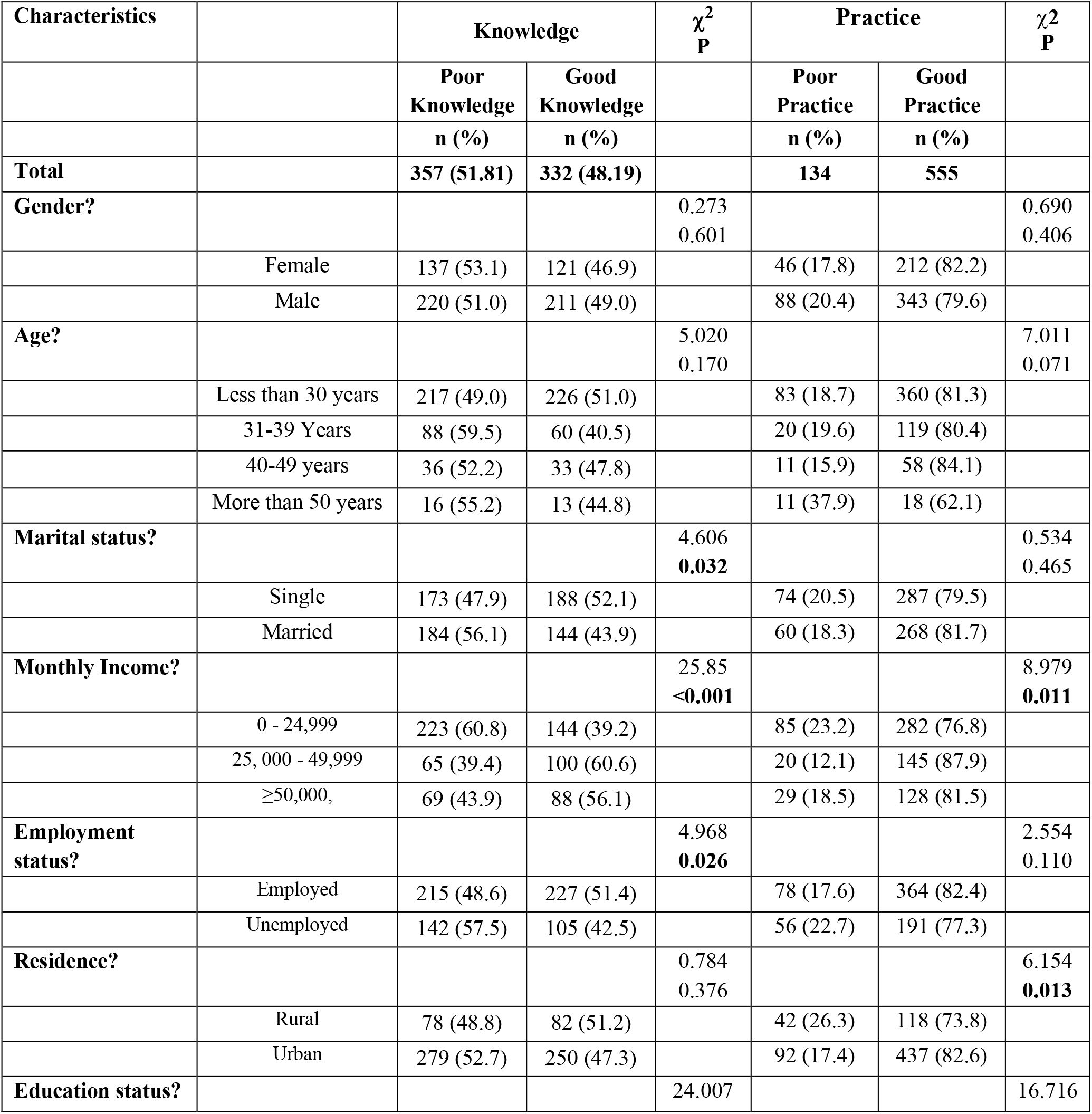
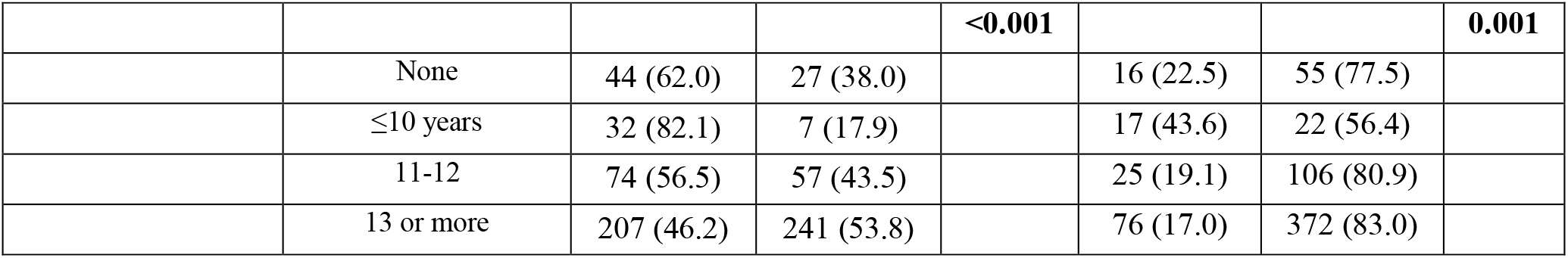
Difference in knowledge and practice status by demographics

**Table 3.**
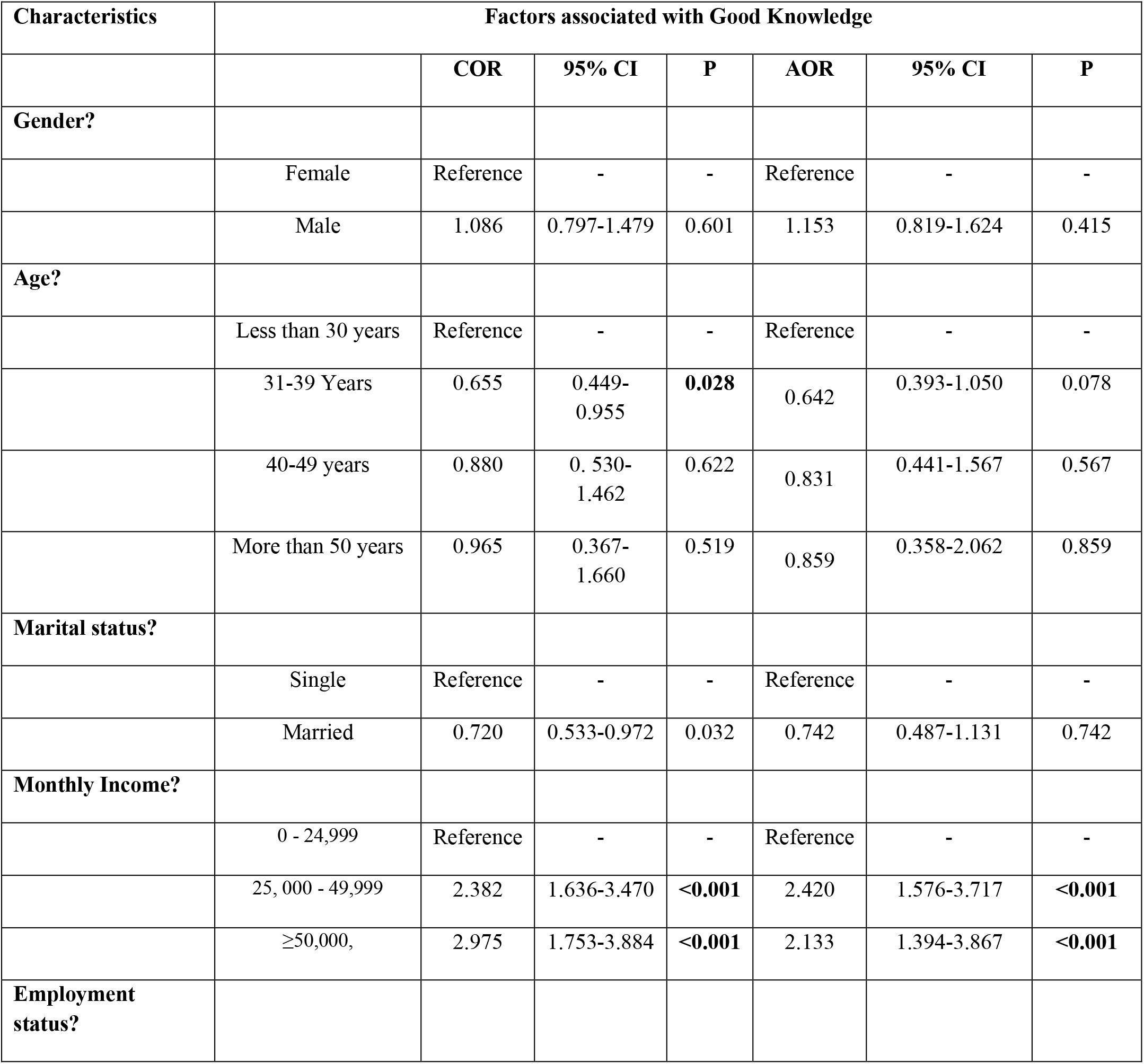
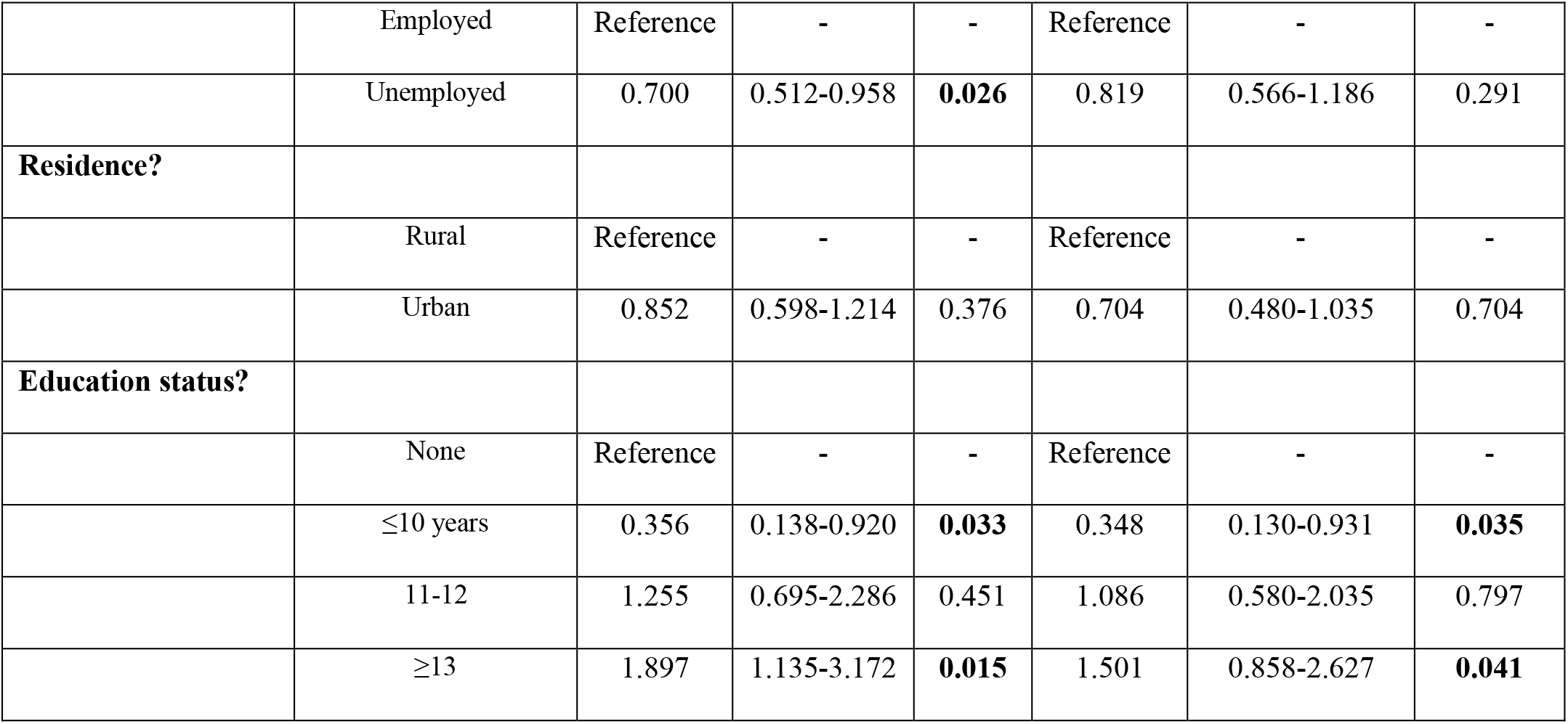
Binary logistic regression analysis for factors associated with good knowledge

| Characteristics | Factors associated with Good Knowledge |  |  |  |  |  |  |
| --- | --- | --- | --- | --- | --- | --- | --- |
|  |  | COR | 95% CI | P | AOR | 95% CI | P |
| **Gender?** |  |  |  |  |  |  |  |
|  | Female | Reference | - | - | Reference | - | - |
|  | Male | 1.086 | 0.797-1.479 | 0.601 | 1.153 | 0.819-1.624 | 0.415 |
| **Age?** |  |  |  |  |  |  |  |
|  | Less than 30 years | Reference | - | - | Reference | - | - |
|  | 31-39 Years | 0.655 | 0.449-0.955 | **0.028** | 0.642 | 0.393-1.050 | 0.078 |
|  | 40-49 years | 0.880 | 0.530-1.462 | 0.622 | 0.831 | 0.441-1.567 | 0.567 |
|  | More than 50 years | 0.965 | 0.367-1.660 | 0.519 | 0.859 | 0.358-2.062 | 0.859 |
| **Marital status?** |  |  |  |  |  |  |  |
|  | Single | Reference | - | - | Reference | - | - |
|  | Married | 0.720 | 0.533-0.972 | 0.032 | 0.742 | 0.487-1.131 | 0.742 |
| **Monthly Income?** |  |  |  |  |  |  |  |
|  | 0 - 24,999 | Reference | - | - | Reference | - | - |
|  | 25, 000 - 49,999 | 2.382 | 1.636-3.470 | **<0.001** | 2.420 | 1.576-3.717 | **<0.001** |
|  | ≥50,000, | 2.975 | 1.753-3.884 | **<0.001** | 2.133 | 1.394-3.867 | **<0.001** |
| **Employment status?** |  |  |  |  |  |  |  |
|  | Employed | Reference | - | - | Reference | - | - |
|  | Unemployed | 0.700 | 0.512-0.958 | **0.026** | 0.819 | 0.566-1.186 | 0.291 |
| **Residence?** |  |  |  |  |  |  |  |
|  | Rural | Reference | - | - | Reference | - | - |
|  | Urban | 0.852 | 0.598-1.214 | 0.376 | 0.704 | 0.480-1.035 | 0.704 |
| **Education status?** |  |  |  |  |  |  |  |
|  | None | Reference | - | - | Reference | - | - |
|  | ≤10 years | 0.356 | 0.138-0.920 | **0.033** | 0.348 | 0.130-0.931 | **0.035** |
|  | 11-12 | 1.255 | 0.695-2.286 | 0.451 | 1.086 | 0.580-2.035 | 0.797 |
|  | ≥13 | 1.897 | 1.135-3.172 | **0.015** | 1.501 | 0.858-2.627 | **0.041** |

Finding showed that age group of 50 years or more (AOR: 1.501, 95% CI: 0.858–2.627, *p* = 0.020), monthly income of PKR 25, 000 – 49,999 (AOR: 1.875, 95% CI: 1.055–3.331, *p* = 0.032), education of 1–10 years (AOR: 1.801, 95% CI: 1.201–2.700, *p* = 0.049), education of 13 years or more (AOR: 1.250, 95% CI: 0.647–2.415, *p* = 0.031), and good knowledge (vs. Poor knowledge: AOR: 1.250, 95% CI: 0.647–2.415, *p* = 0.004) were the substantial determinants of good practice regarding COVID-19 among general population in Pakistan. (Table. 4)

**Table 4.** Binary logistic regression analysis for factors associated with good Practice

| Characteristics | Factors associated with Good Practice |  |  |  |  |  |  |
| --- | --- | --- | --- | --- | --- | --- | --- |
|  |  | COR | 95% CI | P | AOR | 95% CI | P |
| **Gender?** |  |  |  |  |  |  |  |
|  | Female | Reference | - | - | Reference | - | - |
|  | Male | 0.846 | 0.569-1.256 | 0.406 | 0.810 | 0.528-1.1242 | 0.334 |
| **Age?** |  |  |  |  |  |  |  |
|  | Less than 30 years | Reference | - | - | Reference | - | - |
|  | 31-39 Years | 0.946 | 0.591-1.515 | 0.818 | 0.740 | 0.396-1.381 | 0.344 |
|  | 40-49 years | 1.121 | 0.611-2.417 | 0.578 | 0.920 | 0.394-2.145 | 0.847 |
|  | More than 50 years | 1.377 | 1.072-1.829 | **0.015** | 1.087 | 0.428-1.207 | **0.020** |
| **Marital status?** |  |  |  |  |  |  |  |
|  | Single | Reference | - | - | Reference | - | - |
|  | Married | 1.152 | 0.788-1.682 | 0.465 | 1.445 | 0.834-2.503 | 0.189 |
| **Monthly Income?** |  |  |  |  |  |  |  |
|  | 0 - 24,999 | Reference | - | - | Reference | - | - |
|  | 25, 000 - 49,999 | 2.185 | 1.291-3.700 | 0.004 | 1.875 | 1.055-3.331 | **0.032** |
|  | ≥50,000, | 2.330 | 1.831-3.130 | 0.234 | 1.962 | 1.251-2.120 | 0.379 |
| **Employment status?** |  |  |  |  |  |  |  |
|  | Employed | Reference | - | - | Reference | - | - |
|  | Unemployed | 0.731 | 0.497-1.074 | **0.111** | 0.891 | 0.573-1.386 | 0.609 |
| **Residence?** |  |  |  |  |  |  |  |
|  | Rural | Reference | - | - | Reference | - | - |
|  | Urban | 1.691 | 1.113-2.568 | 0.014 | 1.363 | 0.876-2.120 | 0.170 |
| **Education status?** |  |  |  |  |  |  |  |
|  | None | Reference | - | - | Reference | - | - |
|  | ≤10 years | 0.376 | 0.162-0.875 | **0.023** | 0.415 | 0.173-0.995 | **0.049** |
|  | 11-12 | 1.233 | 0.608-2.501 | 0.561 | 1.188 | 0.567-2.489 | 0.649 |
|  | 13 or more | 1.424 | 0.775-2.618 | **0.015** | 1.250 | 0.647-2.415 | **0.031** |
| **Knowledge** |  |  |  |  |  |  |  |
|  | Poor | Reference | - | - | Reference | - | - |
|  | Good | 1.878 | 1.271-2.774 | **0.002** | 1.801 | 1.201- 2.700 | **0.004** |

## Discussion

In view of the rapid spread of COVID-19 and the increase in the number of cases in Pakistan, it is necessary to have a clear picture of the state of public awareness and their practices in the context of the precautionary measures. In addition, Pakistan is a populous country and is facing enormous pressure on non-communicable diseases [17]. Both of these factors increase the country’s vulnerability to this deadly infection and results in higher mortality and morbidity. Moreover, Pakistan’s history of dealing with epidemics required a high level of preparedness by government as well as masses. Global efforts have been made to reduce the transmission of this contagious infection. These efforts include political efforts by the governments, together with personal attitudes and behaviors, which depend on the awareness of the general public about the disease.

Findings revealed that almost half of the population had good knowledge, and 80% had a precautionary approach. News channels such as the internet and social media platforms had become commonly used information sources compared to traditional channels such as newspapers etc. Social media was the primary source of information to be used by the general public (66.62%) in Pakistan to obtain information on COVID-19. This could be explained by the fact that the majority of study participants were under 30 years of age and had university level education. This stratum is the main user of the Internet in Pakistan, according to a recent survey by the Pakistan Telecommunications Authority (PTA), 63% of the 76 million people who have access to the Internet are under 30 years of age [18].

This finding has implications that although social media platforms could be an easily accessible source of information, there is a potential risk of misinformation. As with this pandemic of COVID-19, there is also a pandemic of misinformation on the Internet that leads to negative reactions from the public [19]. Mainly, false information regarding the potential benefits of certain drugs such as hydroxychloroquine stimulated the irrational use of this drug by masses, and this results in a shortage of these medicines and becomes unavailable to patients who need [14].

Results show that 51.81% (n = 357) of the population had good knowledge of the nature, transmission, risk groups and precautionary measures of COVID-19. This rate of good knowledge is lower as reported in a Pakistani study (64.8%) [19], a Malaysian study (80.5%) [20] and a Chinese study (90%) [16]. However, this is in agreement with the findings of Abdelhafiz et al., who reported Egyptians had average (16.39 ± 2.63, range: 7–22) knowledge regarding COVID-19 [14]. A possible reason for less knowledge reported in this study could be explained by the fact that the majority of respondents attained COVID-19 related information from social media. Owing to unauthenticated and the use of social media to get information can explain the existence of myths and misinformation among the general public.

Of note that more than half (55.6%, n = 383) of the study population incorrectly reported that COVID-19 directly leads to death. While wolf et al. reported that only 14.2% of US adults think that COVID-19 may cause death [15]. Possible speculation is that factors such as the outbreak itself and consequential lockdown result in severe psychological impact as khan et al. reported 87.73% of the studied population feared the current situation, which leads to fatigue, anxiety, and depression [21]. Additionally, misinformation surging on the internet and related economic pressure also put immense pressure and creates negative feelings about the situation [22].

The adjusted regression model revealed that higher education and monthly income are substantial predictors of good knowledge (P< 0.005). These results are in line with an Egyptian study which also stated that public awareness was important in terms of its socioeconomic status and level of education (P< 0.002) [14]. A Chinese study concluded that higher education (middle school or lower vs. Master β: 1.346, P< 0.001) played a significant role in increasing good knowledge [16]. While an American study didn’t found any difference in respondents’ knowledge regarding symptoms by poverty level (68.5% vs. 73.1%, P>0.05) [15], this finding is of particular importance for the Government and the authorities concerned to focus on the less privileged stratum of society to ensure the effective implementation of precautionary measures.

Findings indicated that 80% of participants had positive practices in following precautionary measures. This is in line with the results of Zhong et al., who also reported that the majority (>90%) of participants were following precautionary measures [16]. Possible speculation of a higher rate of good practices depsite of only 50% population had good knowledge could be that of campaigns lauched by Government describing causes, symptoms, and route but these awareness campaigns primarily focused on highlighting precautionary measures such as wearing a facemask, social distancing, and hand hygiene practices.

Note that 20.2% of the participants did not wear a face mask when they left their homes. This poroprtion is much higher than the chineese study, which reported that only 2% of the population studied did not wear a face mask. [16]. While Pakistani study reported that 14.2% of the people surveyed did not wear a face mask. Despite the vigorous broadcast of precautionary measures, this risk of taking action could be attributed to the younger age of the participants. Additionally, this might be because face mask shortage in different parts of the country due to high demand as well as price hiking also affects the affordability of the less income stratum. The government had taken several measures to ensure the availability and price control of all personal protective equipment (PPEs) [23].

The adjusted multivariable logistic regression model demonstrated that older age, higher education, and knowledge regarding COVID-19 are the factors that substantially related to the positive attitude among the general public in Pakistan. Zhong et al. found that knowledge was significantly associated with positive practices as Chinese individuals with higher education regarding COVID-19 were less likely to visit crowded places (OR:0.90, p = 0.001) and not wearing a face mask (OR:0.78, p< 0.001) [16]. While a Pakistani study also concluded that younger age (vs. 30 years; OR = 3.08, p< 0.001) and lower education (matriculation vs. Master degree, OR = 6.829, p< 0.001) were the characteristics potentially associated with poor practices regarding COVID-19 [19].

To control the pandemic, there is a need for continuous monitoring of the implementation of preventive measures, the review of existing interventions and the updating of such responses. This study helps to inform the current state of awareness and practices of preventive measures and adds to the findings of a previously conducted study in Pakistan [19].

There are a number of implicit limitations to the study. First, as this is an online survey design, the response depends primarily on honesty and partly on the ability to recall and may, therefore, be subject to bias recall. However, due to a policy of lockdown and social distancing, the survey hand filling was not possible. Second, the sample size is not large enough, and most of the respondents were from one province (Punjab), which limits the generalizability of the population in the whole Pakistan. Potential sample clustering, as the majority (> 60%) of participants were young and students, may also limit the generalizability of the study.

## Conclusion

This quick online survey shows that more than half of Pakistani residents had keen awareness, and 80% had positive practices following precautionary steps. The study is also capable of highlighting gaps in specific aspects of knowledge and practice that should be addressed in future awareness and education campaigns. Findings have also shown that fewer credible sources are used by the general population, which should be discussed immediately because it eventually affects information and is demonstrated in attitudes and practices. The study suggests that the Ministry of Health supports both the corrective and therapeutic steps of COVID-19, consisting of a better-organized approach to all strata of society, i.e., the less privileged, the elderly and the less educated, to provide a balance of knowledge of COVID-19 and hence successful implementation of the precautionary measures.

## Data Availability

Data will be available on request.

## Acknowledgements

The authors would like to extend heartfelt graciousness to all the participants and teachers who provided support at every step of the research.

## Authors’ contributions

The manuscript idea, concept, writing, and layout was done by MS, MM and AG. MMM and SN provided critical help in writing, statistical and layout designing. AH and ZA provided critical input regarding data analysis at every step of the manuscript writing process. MS, SK and ZA proof read the manuscript and provided input in formulating the draft.

